# A Cross-Sectional Analysis of Disparities in Biopsy Result Disclosure for Patients with Limited English Proficiency

**DOI:** 10.1101/2024.11.14.24317354

**Authors:** Joseph Dan Khoa Nguyen, Erica Mark, Seth Martin, R. Hal Flowers

## Abstract

Patients with limited English proficiency (LEP) receiving care from language-discordant providers experience worse outcomes in healthcare quality, safety, and satisfaction, so optimizing medical care for patients with LEP is an integral topic for quality improvement. At our institution, biopsy results are disclosed via telephone unless discussed during an appointment. In this retrospective cohort study, we aimed to quantify the relative lag time in telephone-based biopsy result disclosure when interpretation services were utilized compared to when they were not. A total of 12,372 patient interactions were analyzed; 250 patients (2.02%) required interpreters, and most of these patients spoke Spanish (196, 78.4%). Other languages spoken included Tagalog, Ukrainian, Serbian, Russian, Punjabi, Mandarin, Kurdish, Farsi, Nepali Arabic, and American Sign Language. The mean delay from provider result release to disclosure for patients who required an interpreter was 55.0 hours compared to 36.6 hours for those who did not, respectively (p<0.001). For those requiring a Spanish interpreter, this delay was 60.2 hours (p<0.001). For non-Spanish interpretation, the delay was 36.6 hours (p=0.99). Patients requiring a Spanish interpreter experienced increased mean wait times for receiving biopsy results compared to those who spoke English. Possible reasons include increased communication burden for clinicians when using interpreters, insufficient provider awareness of delays, and patient socioeconomic factors. Future research may analyze the impact of delayed result disclosure, optimal disclosure methods, and initiatives to reduce disparities in time to result disclosure for LEP patients.

## Background

Patients with limited English proficiency (LEP) receiving care from language-discordant providers are more likely to have worse outcomes in healthcare quality, safety, and satisfaction compared to LEP patients or those fluent in English receiving care from language-concordant providers [1-3]. LEP refers to individuals who speak English “less than very well [4].” In 2013, 25.1 million U.S. residents were considered to have LEP [4]. The Joint Commission’s Sentinel Event Database reveals that communication issues are the most common root cause of serious patient safety events [5], so optimizing medical care for patients with LEP is an integral topic for quality improvement.

At our academic medical center, biopsy results are generally disclosed via telephone unless discussed during an appointment. Our institution does have widely used interpretation services that are offered for all patients, both for in-person visits and telephone calls. In this retrospective analysis, we aimed to quantify the relative lag time in telephone-based biopsy result disclosure when interpretation services were utilized compared to when they were not. To the authors’ knowledge, this is the first paper exploring disparities in telephone-based biopsy result disclosure for patients requiring interpretation services.

## Methods

### Participants

This study was deemed exempt by the University of Virginia Institutional Review Board. This study is a retrospective cohort study of patients who received telephone-based biopsy disclosure at an academic medical center. Inclusion criteria were disclosure of biopsy result by telephone, disclosure by a physician, and recorded interpretation status.

### Data Collection

Deidentified patient data from January 2019 - December 2021 were extracted from Epic.

### Measures

Data collected from Epic included time of finalized biopsy reports, time of biopsy report finalization, time of patient receiving results, and interpretation utilization.

### Analysis

Data were exported to R, version 4.0.2. The primary outcome was lag time, defined as the time from when the biopsy report is finalized to when patients receive the results (through phone call). Lag time comparisons were made via Wilcoxon Rank Sum tests between patients requiring interpretation vs no interpretation, and between patients requiring Spanish interpretation vs non-Spanish interpretation vs no interpretation. In this analysis, all patients who utilized interpretation services were included, including patients using an American Sign Language interpreter, to effectively assess the impact of interpretation services on biopsy result disclosure lag time.

## Results

A total of 12,372 patient interactions were analyzed; 250 patients (2.02%) required interpreters, and most of these patients spoke Spanish (196, 78.4%). Other languages spoken included: Tagalog (n=1), Ukrainian (n=2), Serbian (n=2), Russian (n=2), Punjabi (n=1), Mandarin (n=5), Kurdish (n=1), Farsi (n=11), Nepali (n=5), Arabic (n=12), and ASL (n=10). The mean delay from provider result release to disclosure for patients who required an interpreter was 55.0 hours compared to 36.6 hours for those who did not; median delays in disclosure were 23.0 hours and 17.0 hours, respectively (p < 0.001; 95% CI: [-31.4, -9.2]). For those requiring a Spanish interpreter, the mean delay was 60.2 hours (p < 0.001; 95% CI: [-36.7, -10.5]). For non-Spanish interpretation, the mean delay was 36.6 hours (p = 0.989, 95% CI: [(-12.8, 12.9]) (Figure 1).

**Figure 1:**
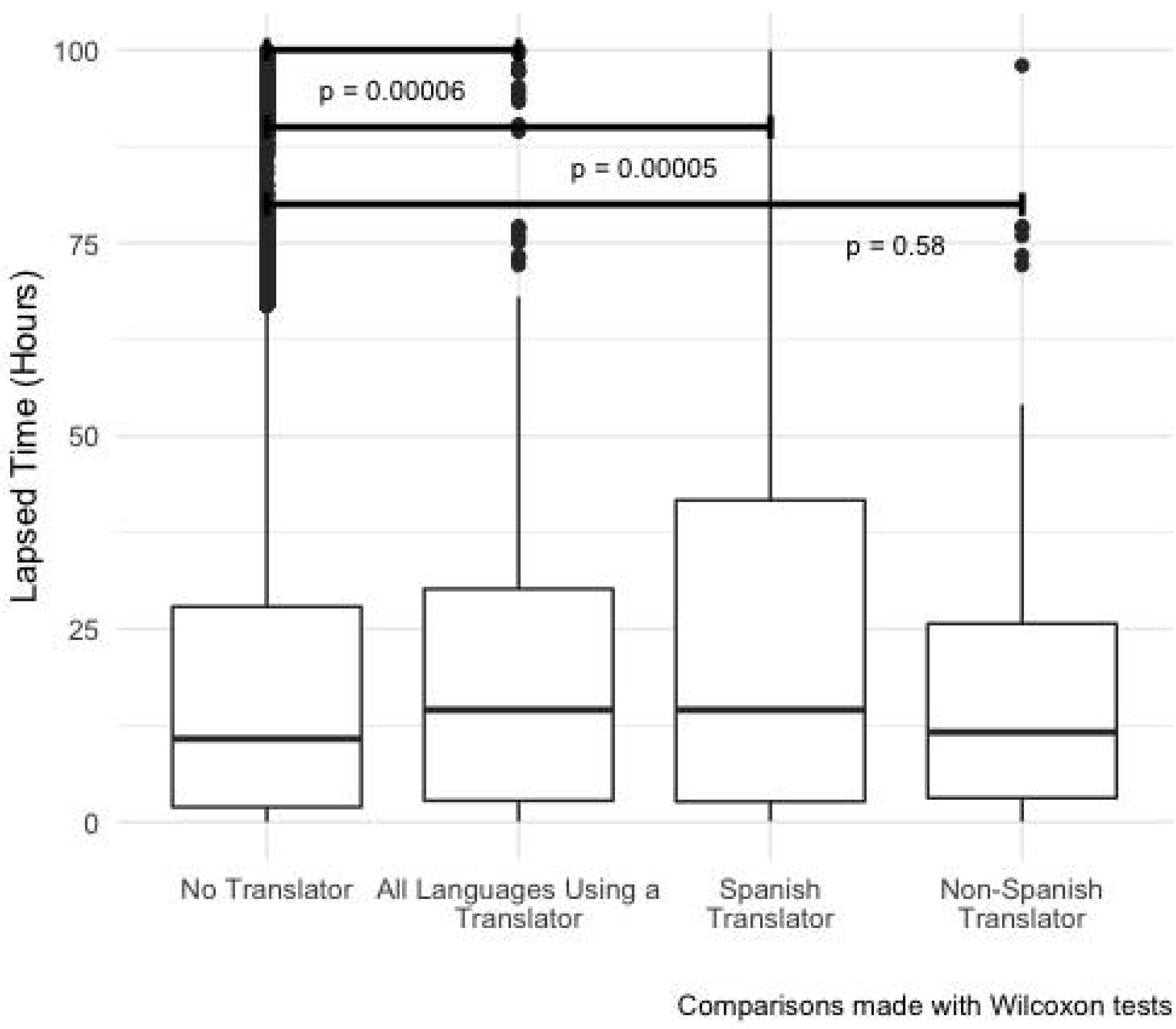
Median and interquartile ranges (IQRs) of lapsed time from provider result release to patient biopsy report disclosure across patients requiring no interpreter, any interpreter, Spanish interpreter, or non-Spanish interpreter.

## Discussion

Based on this data, our study demonstrated a disparity in callback times for patients requiring a Spanish interpreter. Patients requiring a Spanish interpreter had average wait times 23.6 hours (64.4%) longer than those who spoke English or required a non-Spanish interpreter. While receiving a result roughly one day later is unlikely to significantly alter management decisions, the consistent pattern of delays in disclosure of results to patients requiring Spanish interpreters is a notable finding that warrants further investigation. Of note, this delay may lead to delaying future diagnostic and therapeutic procedures, potentially negatively influencing patient outcomes.

Prompt result disclosure to LEP patients requires increased provider time and interpreter availability, creating a communication burden for clinicians [6]. Clinicians report lapses in professional interpretation, communication delays, and feelings of dissatisfaction and ineffectiveness when utilizing a telephone interpreter [7]. Delays in receiving medical results can be stressful for patients [7]. Increased provider awareness of the tendency to inadvertently or advertently delay these call-backs may help them take steps to alleviate the discrepancy in their own practice. While it is plausible that the logistical barriers in using interpretation services may contribute to the delay in call-backs, this does not explain why it only affects patients requiring Spanish interpreters. One plausible explanation for this phenomenon is socioeconomic factors, such as the inability to pay telephone bills or answer calls during work hours, may limit access to effective and timely telephone-based communication. Additionally, the sample size of patients requiring non-Spanish interpretation was very small and may not accurately reflect communication delays in this population. Future studies may be helpful in further characterizing and defining this disparity.

Limitations of this study include: 1) a lack of information on specific diagnoses, as certain diagnoses (e.g. malignancy) may be prioritized over others, leading to shorter callback times, 2) data was from a single center, making it difficult to extrapolate findings across multiple clinical settings, and 3) relatively few patients (∼2%) required an interpreter. Future research may analyze other departments and institutions, the impact of delayed result disclosure, optimal disclosure methods, and initiatives to ensure timely delivery of results for all patients.

### New Contribution to the Literature

To the authors’ knowledge, this is the first paper exploring disparities in telephone-based biopsy result disclosure for patients requiring an interpreter. This paper illuminates a disparity in callback times for patients requiring a Spanish interpreter, which warrants further investigation.

## Data Availability

All data produced in the present study are available upon reasonable request to the authors.

## Notes

### Competing Interest Statement

The authors have declared no competing interest.

### Funding Statement

This study did not receive any funding.

### Author Declarations

This study was deemed exempt by the University of Virginia Institutional Review Board.

